# Estimating the cure rate and case fatality rate of the ongoing epidemic COVID-19

**DOI:** 10.1101/2020.02.18.20024513

**Authors:** Ying Diao, Xiaoyun Liu, Tao Wang, Xiaofei Zeng, Chen Dong, Yuanming Zhang, Changlong Zhou, Xuan She, Dingfu Liu, Zhongli Hu

## Abstract

The epidemic caused by the novel coronavirus COVID-19 in Wuhan at the end of 2019 has become an urgent public event of worldwide concern. However, due to the changing data of the epidemic, there is no scientific estimate of the cure rate and case fatality rate of the epidemic. This study proposes a method to estimate the cure rate and case fatality rate of COVID-19. The ratio of cumulative discharges on a given day to the sum of cumulative discharges on a given day and cumulative deaths before j days is used to estimate the cure rate. Moreover, the case fatality ratio can also be estimated. After simulation calculations, j is statistically appropriate when it is 8-10, and it is also clinically appropriate. When j is 9, based on the available data, it is inferred that the cure rate of this epidemic is about 93% and the case fatality rate is about 7%. This method of estimating the cure rate can be used to evaluate the effectiveness of treatment in different medical schemes and different regions, and has great value and significance for decision-making in the epidemic.

## Introduction

In December 2019, an epidemic caused by the new coronavirus COVID-19 occurred in Wuhan, China. The number of infected people and the spread of the virus have continued to increase, putting pressure not only on China but also the public health security worldwide (1). The rapid research on COVID-19 virology, epidemiology, and clinical medicine has played a positive role in the prevention and treatment of the disease (2-11). Considering the similarity of coronaviruses, the public habitually compares COVID-19 with SARS, especially regarding the data on cure rate (p) (CR) or case fatality rate (q) (CFR). In 2003, SARS emerged. The cumulative number of SARS diagnoses in Mainland China was 5,327, and the cumulative death toll was 349, with a CFR of 6.5 %; meanwhile, its cumulative number globally was 8,096, and the cumulative death toll was 744, with a CFR of 9.6 % (12). However, “there is currently no clear formula for the CR of patients with new pneumonia,” said by Jiao Yahui, the deputy director of the National Health and Medical Commission’s Medical Affairs and Hospital Administration Bureau (13). The popular method of calculating the ratio of cumulative deaths to cumulative diagnosed cases to determine the CFR is accurate to confirm that an epidemic has ended, such as in the case of SARS. However, this estimation method has major flaws when applied in COVID-19, resulting in wrong judgments made by decision-makers regarding its future situation. For example, up to 24:00 on February 16, 2020,the National Health and Medical Commission reported a total of 70,548 confirmed cases, 10,844 discharged patients, and 1,770 deaths. The CR was 15.37 %, and the CFR was 2.51 %. Furthermore, the hospital treatment rate was as high as 82.12 %. Clearly, the current CR and CFR is not the true CR and CFR caused by NOVID-19, nor it can accurately represent the current treatment trend of such a large number of hospitalized patients; if the cumulative discharge and the cumulative death are directly calculated, the CR and CFR would be 85.97 % and 14.03 %, respectively, which also cannot provide enough useful information. Therefore, we designed a method to calculate the CR of the new coronary pneumonia and then estimated the current CR and CFR to provide reference for the diagnosis, treatment, and control decision of this epidemic disease.

## Method

### Data sources

Data were acquired from various provinces, municipalities, and the country from January 20 to February 14. These data included the number of confirmed diagnoses, deaths, and discharges according to the National Health Commission of Wuhan, the National Health Commission of Hubei Province, and the COVID-19 Working Group of the State Council. However, few missing data from January 20 to January 23 were noted. The supplementary principle of the missing data is as follows: when the intermediate data are missing, the average of the two data (before and after) fills the missing data; when the data of Wuhan and the national data are missing, the data of Hubei Province are used as replacement (see Supplement Table S1).

### Defined and corrected estimations of CR and CFR

“Cure” is the end of treatment and the end of death. If the epidemic has ended and all the patients have been treated, the cumulative number of discharged patients is ∑*x*_*i*_, and the cumulative number of deceased patients is ∑*y*_*i*_. When the cumulative number of diagnosed patients is ∑(*x* _*i*_+∑*y* _*i*_), then the CR 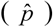 and CFR 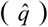 are estimated as follows correspondingly:

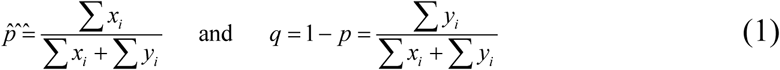

Considering the occurrence of the epidemic situation of the novel pneumonia, some of patients have been cured and discharged, while some are still sick, some died, and some were still under hospital treatment. The insight into whether the patients in the hospital will be cured or die remains unclear. Therefore, speculating the future trend while the epidemic is still ongoing by using the abovementioned formula to calculate the CR or the CFR directly is inappropriate.

According to the results of clinical studies, the novel pneumonia is a self-limiting disease with mild early symptoms, which may worsen after a week, and after the most dangerous period of time, the patient may recover gradually. According to the treatment plan, the patient must wait for all the symptoms to disappear, with two negative results of COVID-19 nucleic acid test, and be isolated for several days before discharge. In other words, the daily reported deceased patients were not the same batch as discharged patients. Hence, confirmers of the same period should consider that a time difference (j day) exists between the cured and death cases.

Therefore, the estimated CR 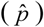 should be the ratio of the cumulative number of discharges on the *i* thday (∑ *x*_*i*_) to the sum of ∑ *x*_*i*_ and ∑ *y*_*i* - *J*_, expressed as follows:

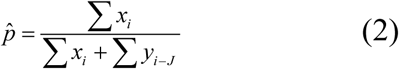

where *y*_*i* - *J*_ is the number of cumulative deaths on the (*i*-*j*)th day and *j* is the parameter that needs to be estimated. The estimated CFR 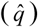 should be the ratio of ∑ *y*_*i* - *J*_ to ∑(*x*_*i*_ + *y*_*i* - *J*_), expressed as follows:

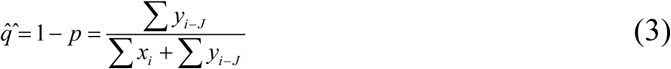

### Determination of the empirical value *j*

The empirical value *j* needs to be estimated before the estimation of the CR and CFR. In statistics, if we do not consider the discovery of specific drugs and medical means in the near future, the best estimates for each rate should be very close to its true value. In other words, these daily estimates are stable, and their variation is minimal. Thus, in this study, the empirical value *j* was determined according to the minimum variance (or coefficient of variation) among the daily estimates.

## Results

### j value

Because of the change in diagnostic criteria on February 12, resulting in a surge in cases, only pre-February 11 data were selected. Taking the national data as an example, starting from February 11 and backtracking, in Equation (2), *j* = 1, 2, 3, …, 15 was calculated, and the CR was calculated daily. When *j* = 10, the value of CR was the most stable among the days, and the coefficient of variation was the smallest.

According to the published data, in addition to calculating the national CR, the same calculations were performed according to the division data of Wuhan, Hubei, Hubei except Wuhan, and the country except Hubei (Fig.1). The coefficient of variation of the estimated cure rate of wuhan, national, Hubei except Wuhan, Hubei and country except Hubei reached the minimum value at j =8,10,10,10,12, respectively, indicating that this is the smallest difference in the estimated value of each day.

**Figure 1.**
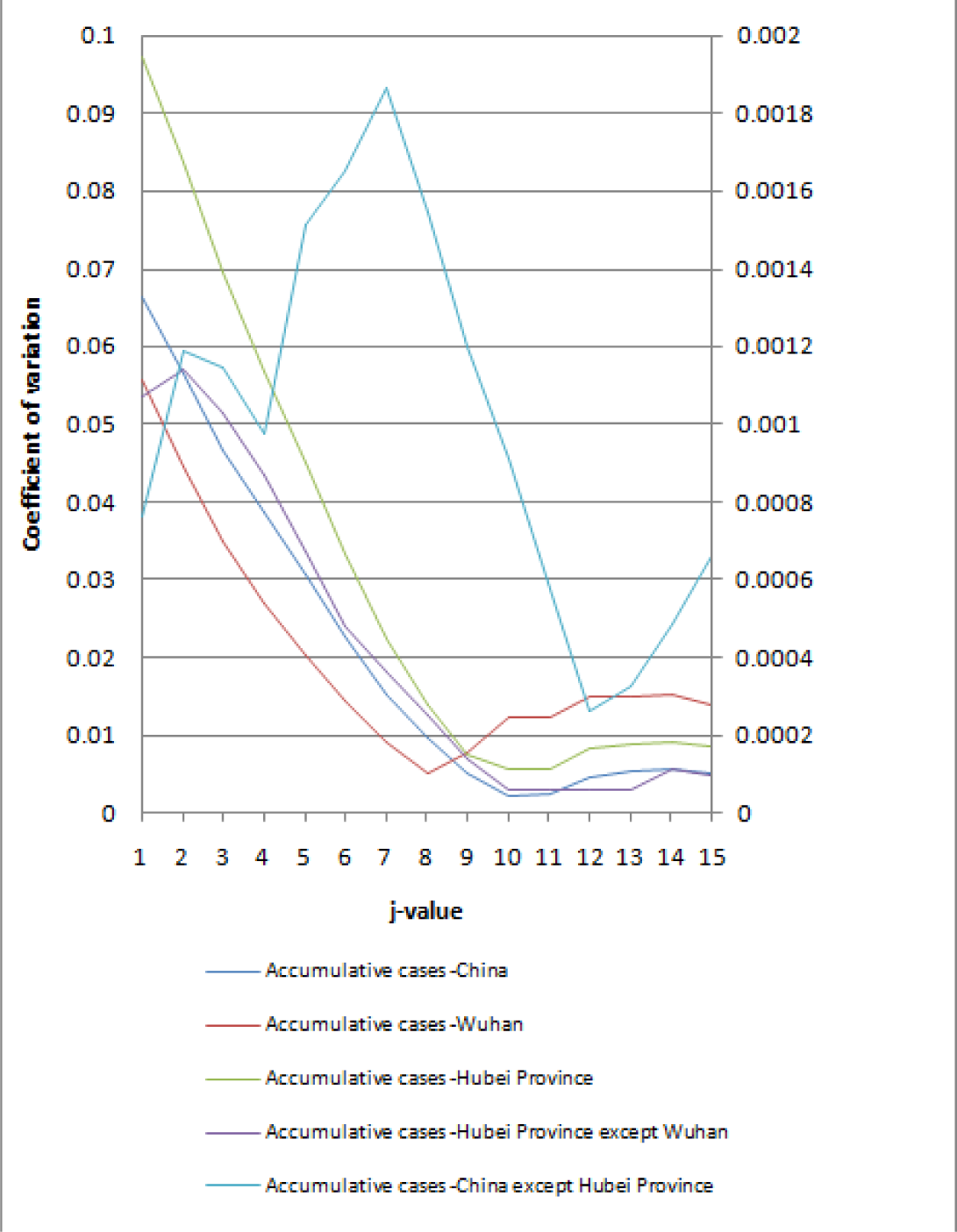
Coefficient of variation of estimated cure rate (p) at different values of j.

### CR and CFR

According to the published data, *j* = 8, 9, 10 were selected. As shown in Fig. 2, the day-to-day CR from February 16 was calculated according to the division of the country, Wuhan, Hubei, Hubei except Wuhan, and the country except Hubei (see Table S2-4). The average CR is shown in Table 1.

**Table 1.**
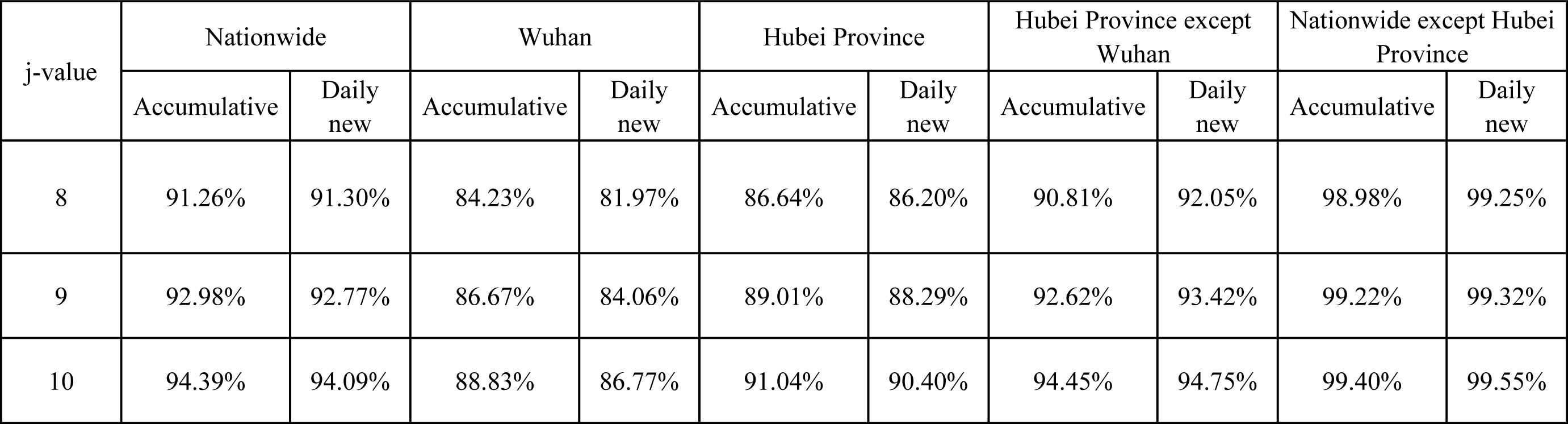
Average of estimated cure rates.

| j-value | Nationwide |  | Wuhan |  | Hubei Province |  | Hubei Province except Wuhan |  | Nationwide except Hubei Province |  |
| --- | --- | --- | --- | --- | --- | --- | --- | --- | --- | --- |
|  | Accumulative | Daily new | Accumulative | Daily new | Accumulative | Daily new | Accumulative | Daily new | Accumulative | Daily new |
| 8 | 91.26% | 91.30% | 84.23% | 81.97% | 86.64% | 86.20% | 90.81% | 92.05% | 98.98% | 99.25% |
| 9 | 92.98% | 92.77% | 86.67% | 84.06% | 89.01% | 88.29% | 92.62% | 93.42% | 99.22% | 99.32% |
| 10 | 94.39% | 94.09% | 88.83% | 86.77% | 91.04% | 90.40% | 94.45% | 94.75% | 99.40% | 99.55% |

**Figure 2.**
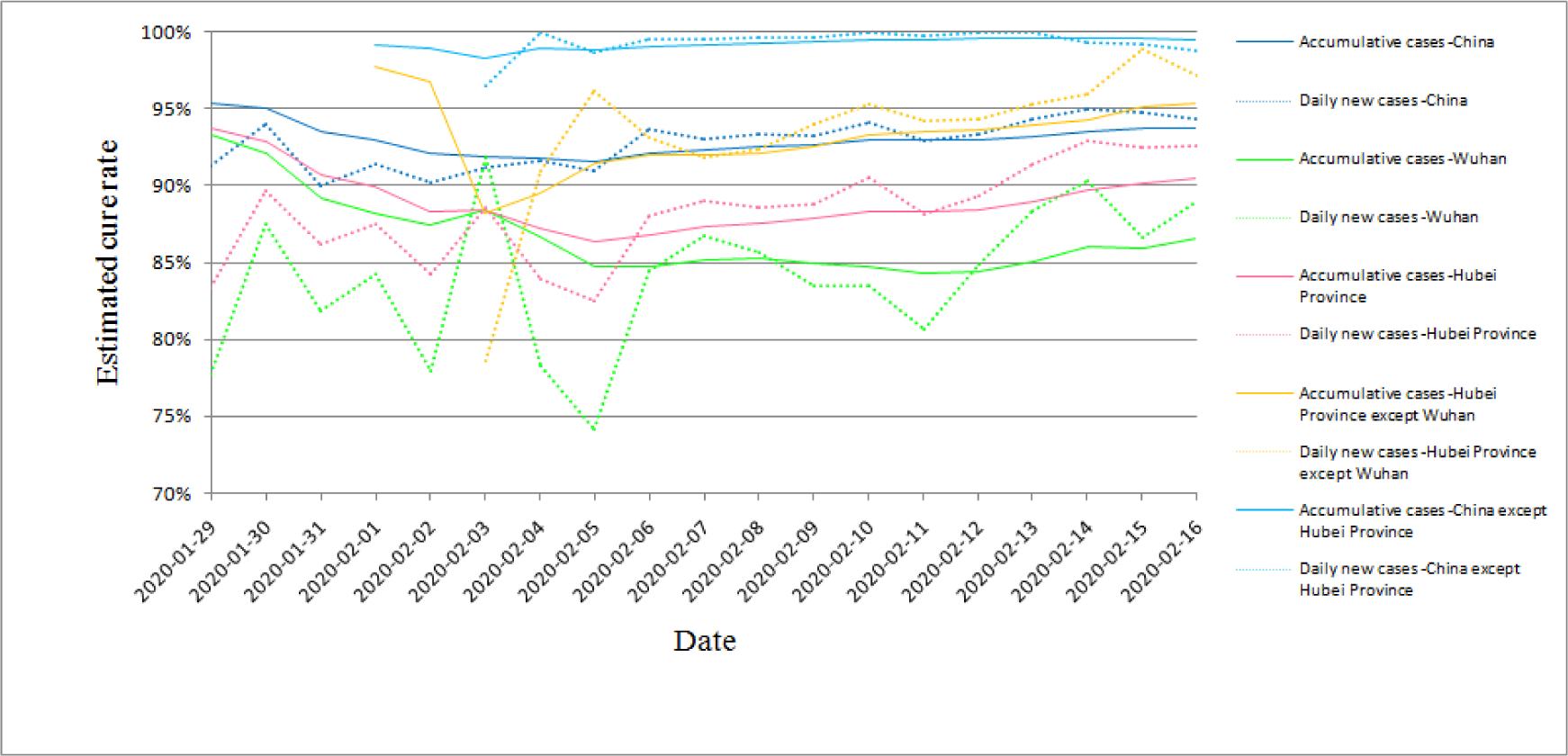
Estimated cure rate at j = 9.

According to Table 1, when *j*=9, if no specific drugs and better treatments emerge later and the pathogenicity of NOVID-19 is not significantly different, the CR of the new coronary pneumonia is approximately 93 %, while its CFR is approximately 7 %, which is relatively close to the CFR of SARS (6.6 %) in Mainland China. In Wuhan, the CR of COVID-19 is approximately 90 %, while its CFR is approximately 10 % or more.

Considering the large number of cases and the low CR in Wuhan, the CR in Hubei Province is lower than the national average, with only approximately 90%. The results of Hubei except Wuhan are similar to the national average. The CR in Hubei is the highest in the country, possibly related to factors such as the large number of imported cases, the different age structure of imported cases, relatively few patients, and better treatment conditions in provinces outside Hubei.

## Discussion

### *j* value

COVID-19 happens all of a sudden, with no known cure. Hence, no treatment experience has been recorded, and no special drugs are currently known. It is mainly managed by symptomatic support treatment, all relying on enhanced immunity.

Assuming that the COVID-19 epidemic event satisfies the statistical randomness, the virus has no variation, and the daily CR and CFR should be basically stable, with approximately the same medical means and measures in a certain period of time and the estimated value of *j* = 9 able to meet this condition.

According to clinical experience, the time point of exacerbation was more concentrated on the 9th to 12th day of the disease course. After the exacerbation of the patient’s condition, a rapid progress process takes place, and death easily occurs (14). In this article, *j* = 9 was consistent with the development of the disease course. Furthermore, according to the data released by the National Health and Medical Commission’s press conference on February 4, the average length of hospitalization for discharged patients in country except Hubei is more than 9 days, whereas that in Hubei Province is 20 days (13). In this article, *j*= 9 was consistent with the actual treatment interval of the disease.

The CR estimate positively correlated with the *j* value size. The larger the *j* value, the greater the CR estimates; the smaller the *j* value, the smaller the CR estimates. In relation to the estimated value of CR, CFR will vary with the value of *j*. Therefore, to obtain accurate estimates of CR and CFR, we need to choose a *j* value, which is also worthy of further study.

However, using this strategy and method is certainly more reasonable than directly “calculating the CR and CFR for the COVID-19 in the country with the ratio of cumulative discharge (∑x) and cumulative death (∑y) to cumulative confirmed cases.”

Moreover, Equation (2) is not only appropriate and applicable during the epidemic; it is also an accurate estimate. The result estimated by Equation (2) is the same as that provided by Equation (1) at the end of the epidemic.

### CR and CFR

This study corrects the current problems in calculating the CR of COVID-19. The CR nationwide is estimated to be 93%, and the CR of Wuhan is approximately 87%, which is considerably higher than the current official data. However, the official CR nationwide on February 16 was only 15.37%. The higher CR we estimated not only helps to boost morale in the country against epidemic disease and alleviate the anxiety of the masses, but also provides a more scientific basis for decision-making on epidemic prevention and control.

Meanwhile, the estimated national CFR of COVID-19 obtained in this study is approximately 7%, which is close to SARS. Considering that the epidemic source is Wuhan, the estimated CFR of Wuhan exceeds 10%. According to the clinical analysis data of Wuhan Jin Yin-tan Hospital, the CFR of the first 41 patients was 15% (9, 15), and the CFR of the first 99 cases was 11% (16). Meanwhile, in the clinical characteristic analysis of the Central South Hospital of Wuhan University, the CFR was 4.3 % (17). We further analyzed the data and found that the number of discharges from Tongji Hospital accounted for 68 % of the study cases, compared with 34.1 %. However, a large number of undischarged cases remain uncertain. Therefore, the statistics of CFR in the Central South Hospital have a marked deviation compared with that in the Jin Yin-tan hospital. The rapid increase in the number of patients, inadequate treatment conditions, delayed diagnosis and delayed treatment, and inadequate knowledge of the virus in the early days in Wuhan affected the treatment effect.

If it is predicted according to the estimated national CFR, under the current medical conditions, the number of national deaths caused by COVID-19 is approximately 4,938 (= 70,548 × 7%), wherein most of them are in Wuhan. In essence, this approach is using the estimated CFR of the first 9 days (February 7) to predict the possible deaths of currently diagnosed patients. The epidemic is continuing, and the number of confirmed patients is still increasing. If no major medical change transpires, the predicted value of future deaths will be extremely large. If the prediction is based on the estimated CFR in Wuhan, the number of deaths caused by COVID-19 is even more alarming. Moreover, all undiagnosed deaths caused by COVID-19 are not included.

This article predicts this high CFR and death toll based on the current medical level and existing data. This result reminds us that we cannot be blindly optimistic. Governments at all levels and people from all walks of life must not take it lightly and must grasp the final “window period.” The patients should be separated and receivable and should receive care and treatment in the best way possible. Identifying high-risk personnel in the treated population, strengthening treatment, and minimizing the CFR should be taken into account. In particular, we need to continue to actively provide medical resources to Wuhan and Hubei Province to control the rate of severe illness and reduce the number of deaths. As the pathogenicity of the coronavirus weakens, with the enhancement of medical technology,improvement of medical conditions, and rich experience in treatment, patients with COVID-19 will be treated more effectively. Consequently, the CR will increase, and the CFR will decrease.

## Data Availability

All data included in this study are available upon request by contact with the corresponding author.

